# Elaboration and validation of R-LAST (Right Language Screening Test), a rapid and reliable screening tool to detect cognitive communication disorders in acute right hemisphere stroke

**DOI:** 10.1101/2025.08.08.25333331

**Authors:** Constance Flamand-Roze, Wendy Régnier, Ryad Zerarka, Heather Flowers, Laura Monetta, Edwige Lescieux, Cosmin Alecu, Didier Smadja, Fernando Pico, Bruno Falissard, Nicolas Chausson

**Author notes:** **Corresponding author:** Constance Flamand-Roze ORCID^®^ iD: 000-0002-8378-5477, **Contact information: email:****, Complete address:** Centre Hospitalier du Sud Francilien, Service de Neurologie, 40 Avenue Serge Dassault, 91100 Corbeil-Essonnes. **Authoship and contributorship:** Conception and design: Constance Flamand-Roze (CFR), Wendy Regnier (WR), Heather Flowers (HF), Laura Monetta (LM) Acquisition of datas: CFR, WR, Edwige Lescieux, Cosmin Alecu, Didier Smadja (DS) Analysis and interpretation of datas: CFR, Ryad Zerarka, Bruno Falissard Drafting the manuscript: CFR Revising manuscript for important intellectual content, final approval of the version to be published: HF, LM, DS, Fernando Pico, Nicolas Chausson.

## Abstract

**Background:** The role of the right hemisphere in language use and interpretive abilities has become clearer through studies describing sequelae in patients with right hemisphere stroke (RHS). Between 50% and 78% of stroke survivors with RHS experience communication disorders, a condition recently termed “apragmatism” by a group of experts. Apragmatism can negatively impact individuals’ personal lives and overall quality of life.

**Objective:** To develop and validate a bedside cognitive and communication screening tool for features of apragmatism in prospective patients with right stroke, called the Right Language Screening Test (R-LAST), which is simple, rapid, and suitable for emergency settings.

**Patients and methods:** R-LAST consists of seven subtests and twelve items. We report its internal and external validity and inter-rater reliability. We validated the scale by prospectively administering it to 300 patients admitted to two stroke units with confirmed right stroke, as well as to 94 stabilized patients with and without cognitive-communication disorders, using the MEC B as a reference.

**Results:** Internal validity demonstrated no redundancy with a Pearson coefficient less than 0.8. The internal consistency of the 12 items was questionable, indicated by a Cronbach’s alpha of 0.62. External validation against MEC B revealed a sensitivity of 0.84 and a specificity of 0.82. Inter-rater agreement was nearly perfect (ICC 0.993). The average time needed to complete R-LAST was 3 minutes and 52 seconds.

**Conclusion:** R-LAST could enable rapid and reliable detection of apragmatism in the acute phase of stroke, which could lead to timely referrals to speech-language therapists and ensure timely rehabilitation. This comprehensively validated cognitive-communication disorders rating scale is simple and rapid, making it a useful tool for bedside evaluation of acute stroke patients in routine clinical practice.

**Clinical Trial Registration:** Unique Identifier: NCT03622606 URL: https://clinicaltrials.gov/study/NCT03622606?term=R-LAST&rank=1

## Introduction

Language function is in the left hemisphere in about 95% of right-handed, 70% of left-handed, and 61.5% of ambidextrous persons [1,2,3]. However, both hemispheres contribute to the richness and efficiency of language use [4]. The role of the right hemisphere in language use and interpretive abilities has become clearer from studies describing sequelae in patients with RHS [5,6]. This lesions may impair higher-level language and interpretive mechanisms related to language use and pragmatic functions, and commonly involve contribution from multiple cognitive domains. comprehension and production, inferencing, prosody (linguistic, emotional, musical), semantic fluency, and visual or verbal semantic judgment, often in association with attentional and executive deficits [4, 7–23]. These impairments, reported in 50% to 78% of patients with RHS lesions [13,18], contribute to reduced social engagement and negatively impact quality of life [7,24].

Initially termed as “crossed aphasia” by Bramwell in 1899 [25], right hemisphere language-related deficits are now recognized as more complex than this term suggests. Over time, clinicians and researchers have struggled to adopt a unified terminology to describe these impairments that affect language use in context, but fall outside classical aphasia. Terms such as “higher-level language deficits” and “cognitive-communication disorders” (CCD) have been used to capture communication impairments linked to other cognitive domains, such as attention, memory, visuospatial processing, executive functions, and social cognition, when these interfere with language interpretation and use [26]. Recently, a group of international experts proposed the term “apragmatism” to designate these right hemisphere deficits, following a concept-mapping process [7]. This definition applies in cases of RHS ipsilateral to the dominant hand, in the absence of childhood brain injury, familial left-handedness [27,28], or other confounding factors such as illiteracy, multilingualism, or ideographic native languages [29].

To the listener, apragmatism can render verbal productions as tangential, imprecise, verbose, or even grossly restricted despite the absence of aphasia. Such impressions can result from challenges using language to make inferences, interpret metaphors, apply attentional processes to maintain a topic and process discourse elements relating to coherence and prosody. These deficits are often nuanced and complex for the listener to identify or address readily in interactions compared to those seen after left hemisphere lesions. Patients with apragmatism retain access to the structural elements of language (e.g., semantics and syntax) but struggle to connect them with the communicative context [11]. The subtlety of deficits, even in the acute phase of the stroke, makes apragmatism challenging to diagnose [11,30–32]. Consequently, affected patients may have restricted access to expert consultation and rehabilitation services, compromising their recovery. Unfortunately, usual care practices often lack routine and comprehensive assessment of persons with right-brain injuries [33]. From an evidence-based care perspective, it remains crucial to perform timely systematic screening for RHS especially since they lend themselves to nuanced yet compromising impairments [13]. Current tests for assessing right hemisphere communication disorders (e.g., MEC B protocol in French [34], The Right Hemisphere Language Battery [35], and even the Cognitive linguistic Quick test [36]) are too lengthy to administer as screening tools during the acute phase of a stroke. To our knowledge, there is currently no available screening tool for apragmatism after acute RHS.

Therefore, this study aimed to develop and validate a rapid detection scale for apragmatism, named R-LAST (Right hemisphere Language Screening Test), applying the same model and methodological approach as for LAST [37].

The project received research ethics approval from the Committee for the Protection of Persons (No. 2017/82).

## Methods

### Scale construction

R-LAST was developed as a formal quantitative scale to assess cognitive and communication abilities related to features of apragmatism resulting from RHS, with the goal of creating a brief and clinically applicable screening tool for use in the acute phase of stroke. A panel of experts in speech-language therapy (CFR, WR, HF, LM) proposed items related to common deficits, selecting the final set by consensus. To address any potential administrative or content ambiguities, the scale was trialed with 52 healthy volunteers. All R-LAST items were successfully completed by the healthy volunteers, regardless of their age or education level. However, some instructions required modifications for clarity (Data not shown).

The accepted scale consists of seven subtests and twelve items (Figure 1). The tool involves a single double-sided sheet of black-and-white paper presented in portrait format to accommodate visuospatial deficits and perceptual disorders common in right hemispheric lesions [29,38].

**Figure 1:**
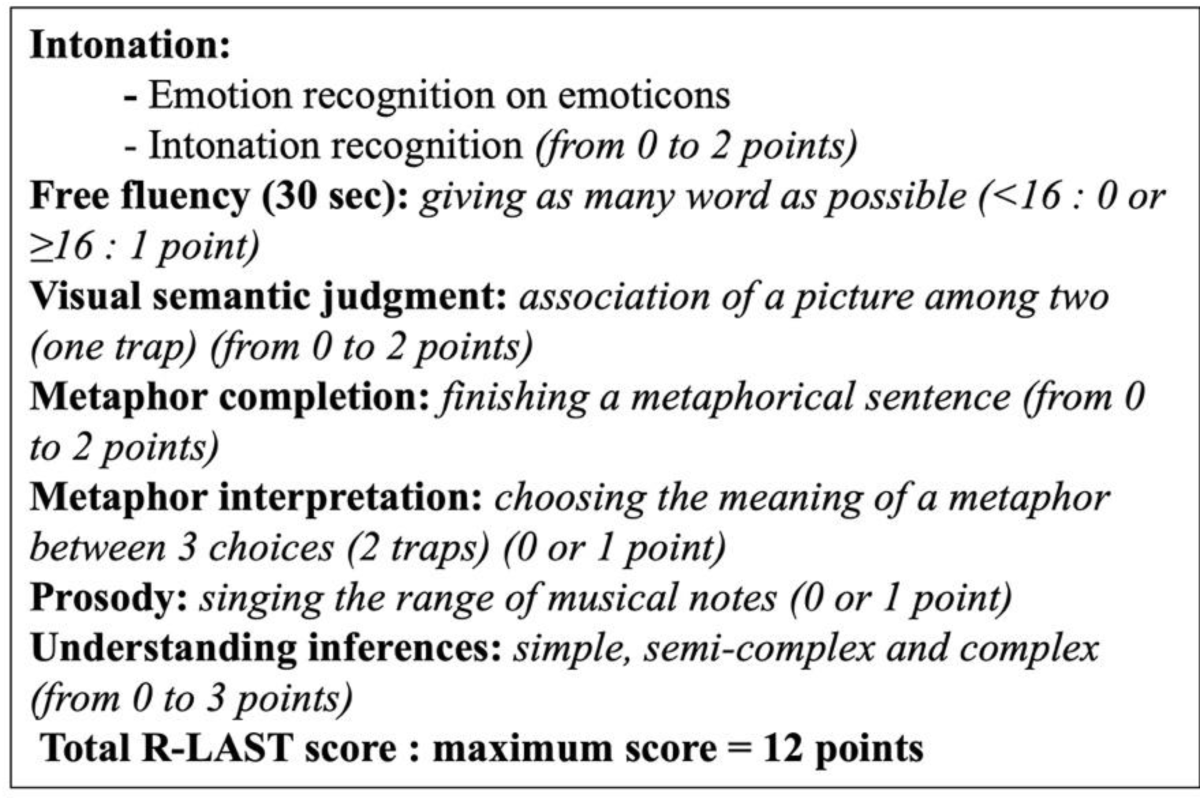
Design of R-LAST (Right Language Screening Test)

The subtests of R-LAST include:

1. Intonation [9,24]; (2) Free fluency [15,16]; (3) Visual semantic judgment [4,9,11,38]; (4)

Metaphor completion [8,10,19]; (5) Metaphor interpretation [19,20,24]; (6) Prosody [8,11,18,24]; (7) Understanding inferences [14,24].(See Supplemental Methods)

### Patients and Instruments

Patients were assessed during the acute phase, from day 0 to day 4 after the onset of symptoms (termed “acute”), for internal validity and inter-rater reliability measures and during the initial post-stroke outpatient consultation occurring at least one-month post-stroke (to ensure compliance and stamina for lengthy testing) for external validity (termed“stabilized”) measures. Not all patients evaluated in the “acute” phase were reassessed during the “stabilized” phase due to medical or logistical constraints.

The study maintained the standard care practicefor hospitalized patients during the acute phase of stroke and those attending post-stroke consultations. The clinical examinations reflected those typically performed in the same contexts, including information about the presenting stroke, the premorbid level of functioning, medical history, NIHSS, LAST, and a laterality questionnaire. (Edinburgh Inventory [39]). The total duration of study-specific assessments comprised the administration of i) R-LAST, ii) the gold standard for external validation (MEC B = 30-45 minutes), and iii) the Edinburgh Inventory (5 minutes). Each participant received written study information prior to inclusion and provided signed informed consent.

### Validation of R-LAST

#### Internal validation

Was conducted from May 2020 to November 2024 in 300 consecutive patients admitted to the acute stroke centers at CHSF (n=150) and CHV (n=150) and who met the inclusion (right hemispheric stroke confirmed by MRI or CT scan, completely right-handed as indicated by the Edinburgh questionnaire, acknowledgment of the information letter, and consent to participate, affiliated with a social security scheme) and exclusion (non-French speakers, stroke history, dementia, sensory impairments including deafness or blindness, family historyof left-handedness, illiterate patients) criteria. Patients who passed LAST were included in the study (score of 15/15), but a single error was permitted (score of 14/15) for the automatic series item, given the known contribution from the right hemisphere for this type of task [40]. That is, an error on this item could correspond to altered prosodic and/or mental organization for automatic speech pathways and should not constitute an exclusion criterion.

#### Inter-rater reliability

Two examiners were present at the bedside: a member of the medical team and a speech-language therapist. One of the two examiners administered R-LAST (as part of the internal validity evaluation); the second examiner also had a copy of R-LAST and recorded the patient’s answers independently remaining blind to the other examiner’s scoring documentation.

#### External validation

Involved 94 patients in the stabilization phase during post-stroke consultations at CHV and CHSF (50 with higher-level language deficits consistent with apragmatism and 44 without) to evaluate R-LAST’s sensitivity and specificity against a gold standard (MEC B). A speech therapist administered MEC B, followed by R-LAST, given by an examiner blinded to MEC B results. Patients were classified as “apragmatic” or “non-apragmatic” based on MEC B results. Administration times for R-LAST and MEC B were recorded. (See Supplemental Methods)

### Statistical Analysis

Internal validity: first, we closely inspected the score distribution for each item to detect a floor or ceiling effect (considered present if more than 90% of respondents achieved a score of 1 or 0). A Pearson correlation matrix was used to detect item redundancy. Second, the number of underlying dimensions was determined through parallel analysis, which involved creating a traditional scree plot alongside simulations [41,42]. Third, we calculated the Cronbach’s α coefficient, a measure of internal consistency based on the correlation of the items within the scale. Inter-rater reliability for total scores was assessed using the intraclass correlation coefficient (ICC). External validity was evaluated in relation to the MEC B. We displayed the Pearson correlations between the R-LAST and MEC B subtests on a sphere [43] and illustrated the Pearson correlation between the total R-LAST and MEC B scores with a scatter plot. We used R 4.1.3 software and the ‘psy’, ‘hms’, ‘mice’, and ‘VIM’ packages for all analyses [44]. The cutoff value was selected to optimize sensitivity, as R-LAST is intended as a screening tool.

## Results

300 patients were included in the assessments of internal validity and interrater reliability (See Figure 2): 171 men and 129 women; mean age 67.6 years (±14.5); 266 ischemic strokes (88,6%), 34 hemorragic strokes (11,4%); mean R-LAST score 8.9 (± 2.0).

**Figure 2:**
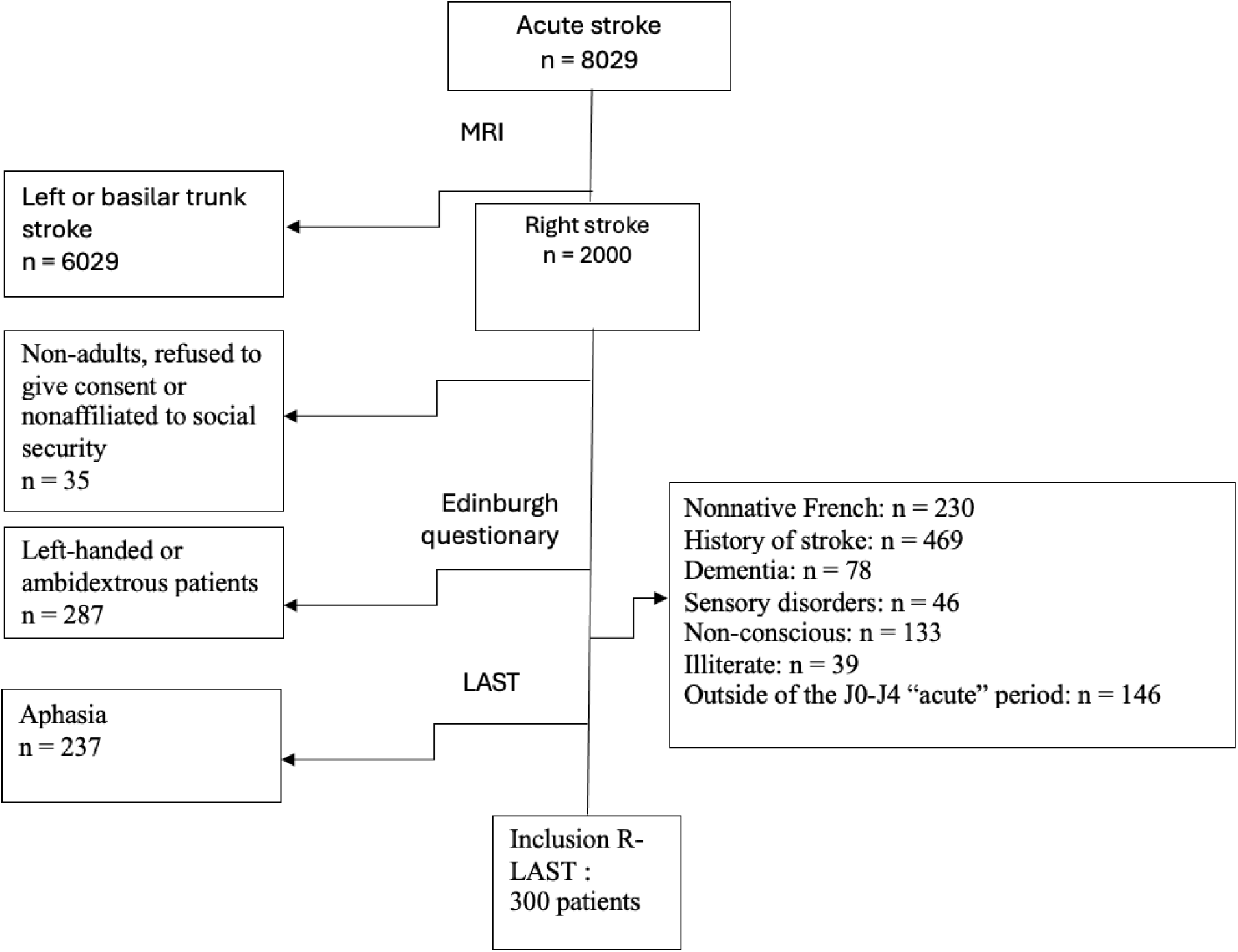
Flow-chart of inclusions and exclusions for “acute” patients

The sample of 94 “stabilized” patients used to evaluate external validity included 58 men and 36 women, averaging 61.9 years (± 16.0) with an average R-LAST score of 10.0 (± 1.5). According to the MEC B results, 50 of these “stabilized” patients (34 men and 16 women, mean age 67.0 years (± 13.3)) had apragmatism, while 44 (24 men and 20 women, mean age 56.1 years (± 17.1)) did not have CCD. (See Figure 3)

**Figure 3:**
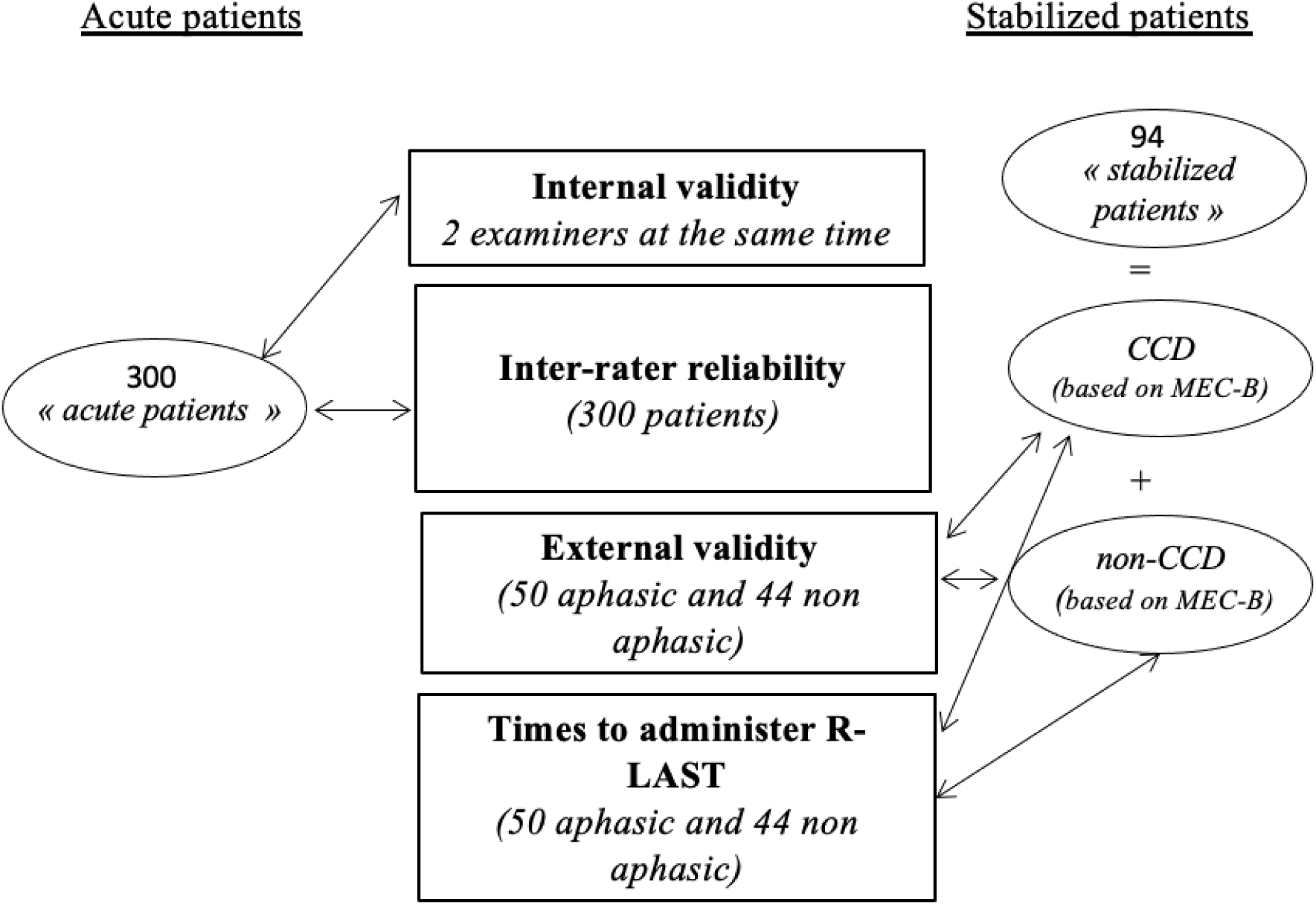
Schematic representation of the R-LAST validation process

### Time Taken to Administer R-LAST

The mean time required to complete R-LAST was 3 minutes and 52 seconds (standard deviation: 27 seconds), whereas completing the MEC B required a mean of 28 minutes and 10 seconds.

### Internal Validity

An item-by-item analysis of the entire sample of 300 “acute” patients revealed ceiling effects on two items (Metaphor interpretation and the first item of the Inferences subtest) and no floor effects. There was no redundancy among items, as indicated by Pearson correlation coefficients less than 0.8 (Figure 4). Parallel analysis revealed a 1-dimensional structure as per shown in the scree plot (See Table 1 in Supplemental Results). The internal consistency of the 12 items was questionable, as indicated by a Cronbach α of 0.62.

**Figure 4:**
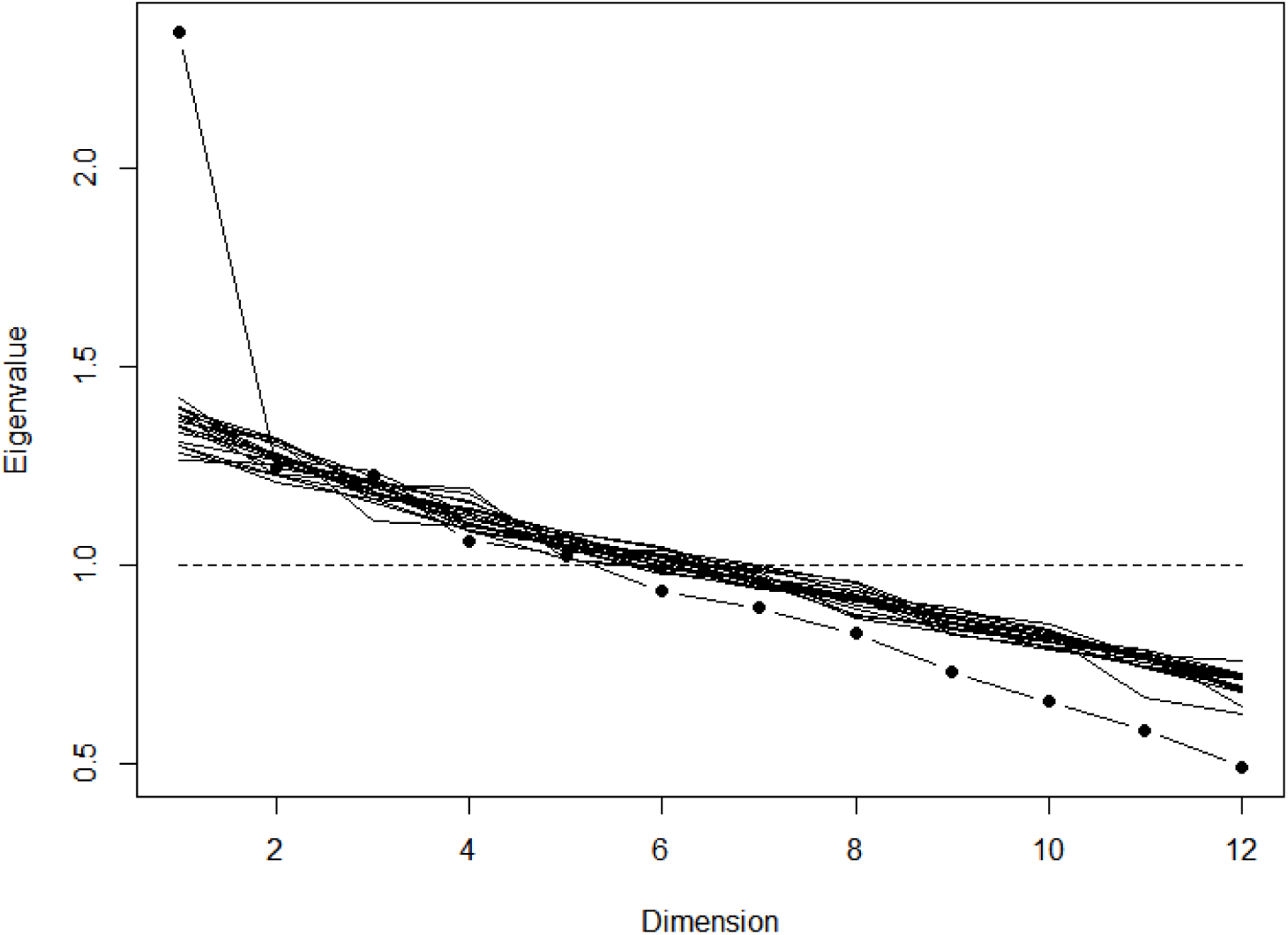
Scree-plot of eigenvalues: Principal component analysis showed a one-dimensional structure

### Interrater Validity

Interrater reliability for the 300 “acute” patients was nearly perfect with an ICC of 0.993.

### External Validity

Patients were tested at an average of 2.1 months after stroke.

A spherical representation of the correlation matrix for the R-LAST and MEC B subtests is depicted to display the strength of correlations (the closer the points are, the stronger the correlation) (See Figure 5). The Pearson correlation calculated between the total R-LAST and MEC B scores was 0.66, as represented by a scatter plot. (See Figure 6)

**Figure 5:**
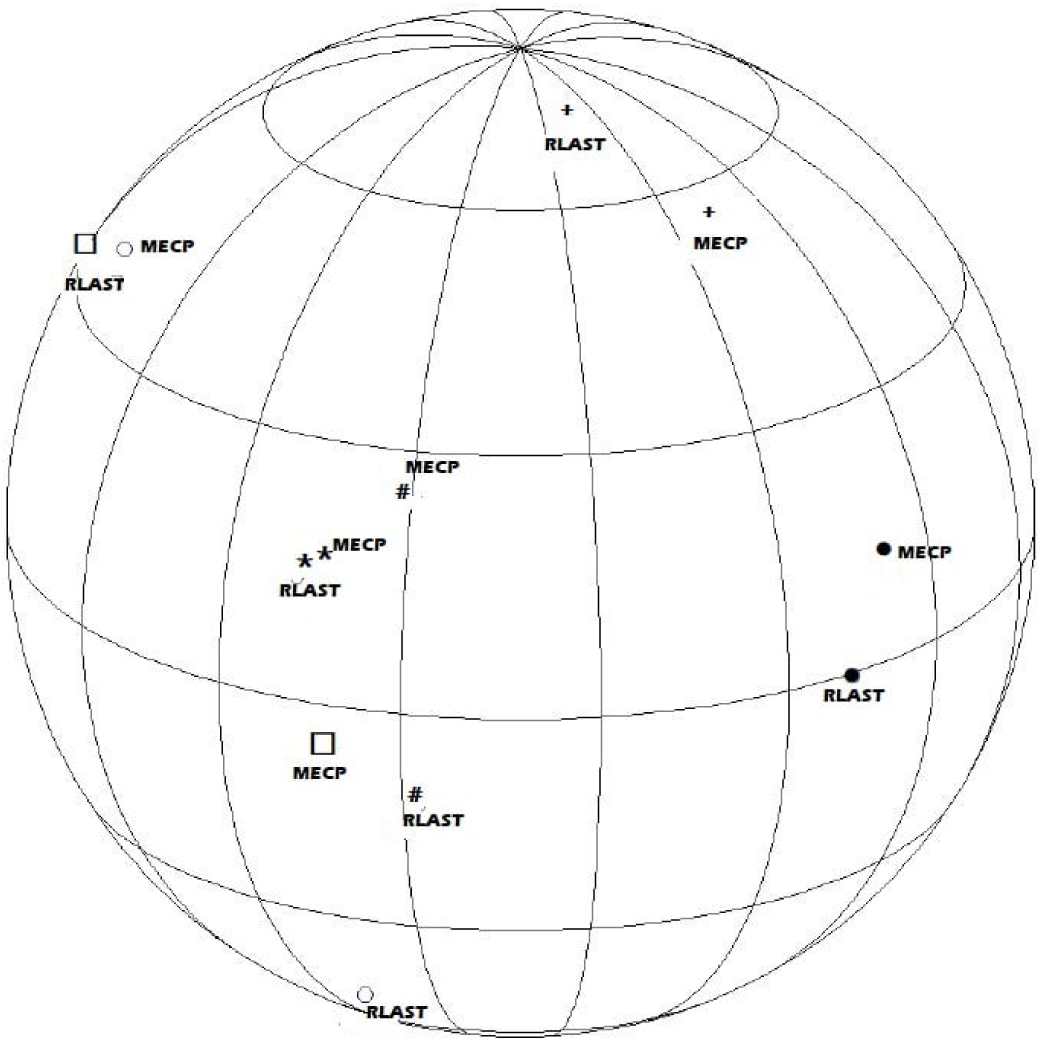
Spherical representation of the correlation matrix for the R-LAST subtests and the corresponding MEC B items. ***+*** *Intonation (R-LAST: 2 items/2 points; MEC B: 2 items/4 points)* *Prosody (R-LAST: 1 item/1 point; MEC B: 5 items/10 points)* *□ Inferences (R-LAST: 3 item/3 points; MEC B: 7 items/12 points)* *\* Fluence (R-LAST: 1 series/1 point; MEC B: 1 series/1 point) # Metaphor (R-LAST: 2 items/2 points; MEC B: 6 items/12 points)* *○ Semantic Judgment (R-LAST: 2 items/2 points; MEC B: 6 items/6 points)* The “metaphor interpretation” item is not represented on the sphere because it is constant.

**Figure 6:**
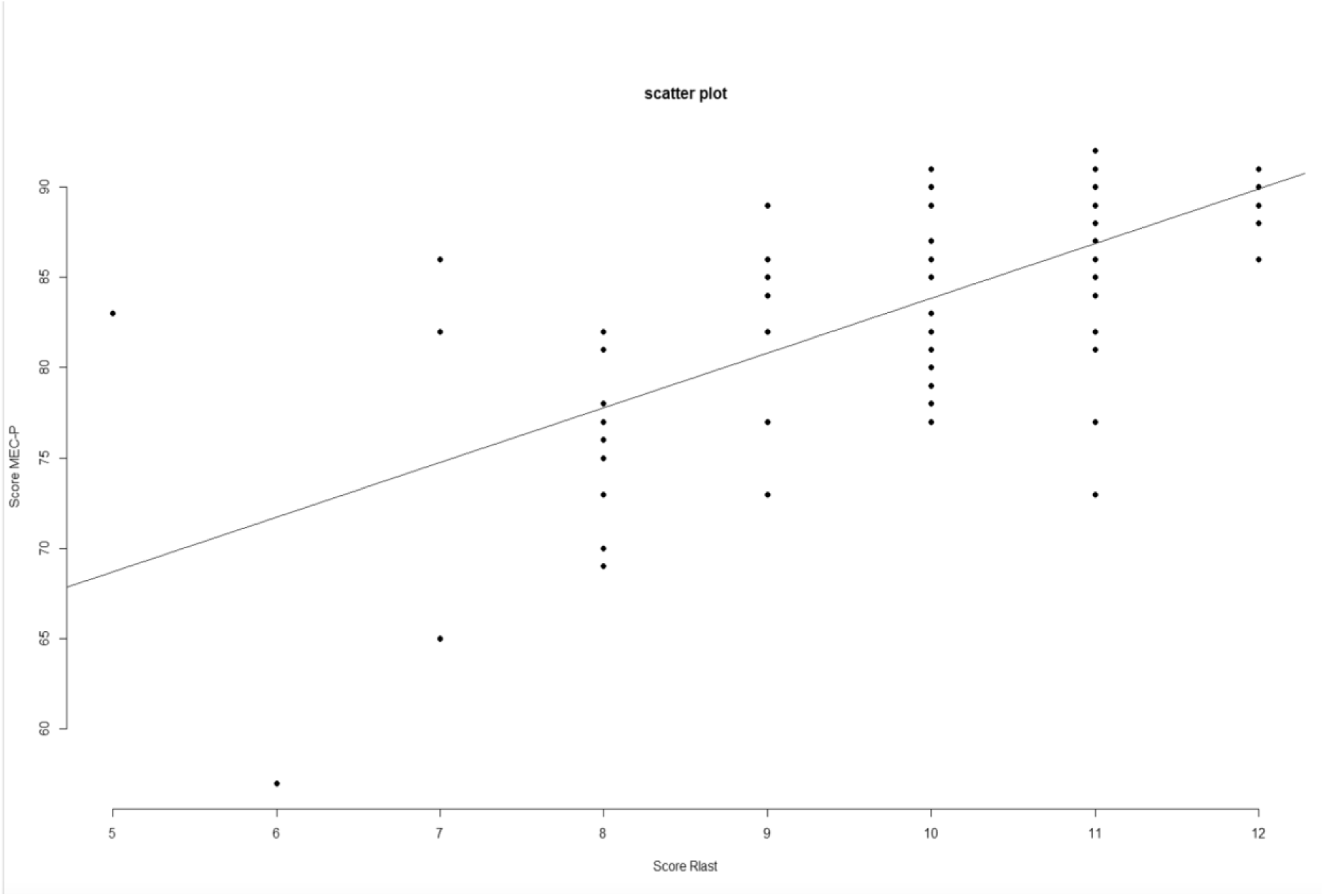
Calculated Pearson correlation between the total scores of R-LAST and MEC B represented by a scatter plot.

R-LAST showed a sensitivity of 84% (95% CI: 71.5–91.7)for apragmatisl, a specificity of 82% (95% CI: 68.0–90.5), a positive predictive value of 84% (95% CI: 71.5–91.7), and a negative predictive value of 81.8% (95% CI: 68.0–90.5), with a cut-off score of 11. Out of the 50 patients identified as “apragmatic” with the MEC B, eight scored 11 or higher in R-LAST (false negatives). Conversely, for the “non-apragmatic”group, eight out of 44 patients scored <11 in R-LAST (false positives).

## Discussion

We developed and validated on a large sample of patients a right hemispheric stroke-specific scale (R-LAST) to screen for apragmatic deficits, predominantly in the realm of oral language and associated interpretive mechanisms, across a range of tasks relating to prosody, non literal language, and affect interpretation in the acute and subacute phases of stroke. The current psychometric analysis has demonstrated strong internal validity and strong correlations with the gold standard MEC B for R-LAST. It also demonstrated excellent inter-rater reliability alongside a rapid administration time, averaging a mere 3 minutes and 52 seconds compared to 28 minutes and 10 seconds for MEC B. Our findings suggest that R-LAST has good diagnostic accuracy for detecting apragmatism in stabilized right hemisphere stroke patients, with sensitivity and specificity above 80% and reasonably narrow confidence intervals (sensitivity: 71.5–91.7%; specificity: 68.0–90.5%). These estimates were obtained in a well-powered validation sample (n = 94), with balanced groups, as recommended by statistical guidance for diagnostic studies.

Our cutoff score of 11 points (from a total of 12), demonstrated excellent sensitivity and specificity for risk of apragmatism in patients with right hemispheric stroke.

– Usefulness in clinical practice: By enabling rapid and effective detection of apragmatism in the acute phase of stroke, R-LAST will precipitate referrals to speech-language therapists and ensure timely rehabilitation for patients who have often remained underidentified due to the nuanced nature of their deficits. It is well established that patients with right hemisphere aprosody are tested less systematically, if at all. Yet, apragmatism can be as equally compromising as aphasia, impacting a range of communication activities required for optimal daily functioning. Indeed, several case reports describe patients for whom apragmatic difficulties persist long-term significantly impacting daily life, with researchers recommending targeted clinical management. [4, 10, 11, 45].

R-LAST detected apragmatism in 79% of the 300 patients admitted to our two stroke units (cutoff score <11/12, as established through external validation), which is in the upper range of what has been described in the literature [11]. This could be attributed to the high sensitivity of R-LAST to apragmatism whereas its frequency in previous studies involved informal evaluations and may have detected only those patients with more severe deficits. By contrast, our patients had a mean score of 8.8/12, well below the cutoff of 11/12 suggesting greater heterogeneity of severity levels. Even mild or moderate deficits can cause severe communication difficulties in daily activities and should deserve recognition. Because R-LAST was administered during the acute phase (between D0 and D4), it included patients who might experience rapid stroke evolution and spontaneous recovery; that is those who may otherwise have been overlooked.

R-LAST is a brief yet reliable test, and can serve as an adjunct to LAST, offering a comprehensive language screening for patients with RHS. Such a tool is badly needed for routine clinical practice, because apragmatism after RHS often presents more covertly than aphasia and may remain undetected during informal bedside examinations. Unfortunately, deficits may emerge when they begin to compromise professional and personal life, pointing to the great need for early management that can propel recovery and mitigated these impacts [10, 11, 45].

In its 2022 guidelines on rehabilitation in the chronic phase of stroke, the HAS (High Authority of Health) in France emphasizes the importance of initiating early and intensive management of patients with communication disorders by speech-language therapists, starting in the acute phase, to maximize functional recovery, in line with other international stroke guidelines [46]. Therefore, timely and routine identification of communication disorders, regardless of lesion laterality, is essential for the swift implementation of rehabilitative care to ensure that the patients receive therapeutic engagement for recovery related to early diagnosis.

– Usefulness for research: R-LAST could enhance our understanding of right hemisphere higher language functions and their relationship to other communication disorders such as aphasia or concomitant cognitive dysfunction. Furthermore, R-LAST could help us better understand the neuroanatomical mechanisms underlying recovery patterns in patients with apragmatism and provide novel insight into the right hemisphere’s communication capacities.

– Usefulness for medical and paramedical fields education: R-LAST could help raise awareness among future neurologists, speech-therapists, and patient caregivers about apragmatism stemming from RHS Our study had several limitations:

– In the development of our scale: Although reading disorders have been identified after RHS, their manifestation is much less frequent than after left hemisphere stroke [47]. We, therefore, decided not to include our initial reading items in R-LAST because of a probable ceiling effect, and this also facilitated a shortened the administration time of the tool. Similarly, we eliminated a “verbal semantic judgment” subtest and prioritized visual semantic judgment, as a more frequent source of deficit [4,27]. Therefore, during external validation, we compared our “visual semantic judgment” subtest with the “verbal semantic judgment” of the MEC B, which does not include a visual modality subtest. While this comparison relied on different input channels, we considered it acceptable given that both tasks assess higher-order semantic integration, regardless of the modality. Furthermore, only 10% of patients with apragmatism scored outside of normal limits on the “verbal semantic judgment” subtest of the MEC B. In comparison, 30% failed the “visual semantic judgment” subtest of R-LAST, suggesting that the visual modality may be more sensitive to right hemisphere deficits.

We also decided not to include a conversation, narrative retell, or picture description as tasks due to administrative time. Although these tasks could enhance sensitivity, they were unnecessary given our strong accuracy statistics.

Two items exhibited a near ceiling effect during the internal validation: only 2% of patients failed the metaphor interpretation item, and 4% failed the first item of the inference subtest. This raises the question of whether to remove these items. During external validation, the MEC B metaphor comprehension items were failed more frequently than those on the R-LAST. This might indicate that additional items should have been added to R-LAST to better identify deficits in metaphor comprehension. Nevertheless, the sensitivity and specificity of R-LAST remained compelling in detecting higher verbal language skills consistent with apragmatism, despite the limited number of items. We advocate for an accurate yet rapid tool that can promote routine implementation. We maintain that both the metaphor interpretation and the first item of the inference subtest should be maintained, particularly since the latter item can serve as practice for the remaining items in the subtest. The sensitivity and specificity largely remained unchanged when we excluded the two items from the analysis.

– Regarding the validation of the scale: we chose to conduct external validation on patients who were at least one month post stroke because (1) there was no other validated and standardized test for right hemisphere communication deficits to compare with R-LAST, and (2) the MEC B was too lengthy to administer in the acute phase of stroke. We validated R-LAST against MEC B based on age and education level by weighting the R-LAST scores to what was pathological in the MEC B ratings for age and level of education.

Given its reliability and brief administration time, R-LAST could support future research, particularly as a thrombolysis decision-making aid in acute stroke settings similar to the use of LAST in thrombolysis protocols [48]. This is relevant since patients with apragmatism are at risk of disabling outcomes and may benefit from timely reperfusion therapies. Longitudinal studies could explore R-LAST’s prognostic value and responsiveness to change. In parallel, lesion-symptom mapping could help identify the neuroanatomical substrates of specific deficits. Finally, we advocate for culturally informed adaptations of R-LAST, rather than direct translations, as was done for LAST [49]. Notably, harmonization of the English version was initiated early to support international use.

## Data Availability

All data referred to in this manuscript are available from the corresponding author upon reasonable request.

## Acknowledgments

None

## Sources of funding

None

## Disclosures

None

## Notes

### Competing Interest Statement

The authors have declared no competing interest.

### Clinical Trial

Clinical Trial Registration: Unique Identifier: NCT03622606

### Funding Statement

No external funding was received.

### Author Declarations

Comité de Protection des Personnes Nord Ouest II Adresse: Bâtiment Pharmacie - Hôpital Nord - Place Victor Pauchet 80054 AMIENS France Courriel: Téléphone: 0322668543

